# Evaluating a Sexual Violence Primary Prevention program in Australian Secondary Schools: A Protocol for a Pilot Cluster Randomised Controlled Trial

**DOI:** 10.64898/2026.05.14.26353223

**Authors:** Freda Haylett, Jacqueline Kuruppu, Jessica Ison, Jacqui Theobald, Gabriel Caluzzi, Xia Li, Innocent Mwatsiya, Sarah Vrankovich, Kerryn O’Rourke, Adam Bourne, Kirsty Forsdike, Nicola Henry, Felicity Young, Leesa Hooker

## Abstract

**Background:** Sexual violence is a global human rights issue and a significant public health concern. Prevention of sexual violence requires addressing the structural and social norms that perpetuate it. Schools are promising settings for primary prevention interventions, as early socialisation experiences can shape attitudes and behaviours that enable sexual violence. However, evidence on effective school-based interventions is limited. The objective of this pilot study is to assess the feasibility and preliminary effectiveness of an Australian sexual violence primary prevention program, the Schools Education Program, on student sexual violence knowledge, attitudes and behavioural intentions.

**Methods/design:** A two-arm, parallel pilot cluster randomised controlled trial will be conducted with Year 9 students (aged 13-15) in 12 secondary schools across one Australian state. Ten schools will be randomly allocated to the SEP intervention (n=6) or a waitlist control (n=4); an additional two schools are pre-assigned to the waitlist control group. The SEP comprises three student education modules, parent and staff education, and the recruitment of School Student Champions. The primary outcome is sexual violence knowledge. Secondary outcomes include attitudes, behavioural intentions, and implementation factors. Quantitative data will be collected at baseline, immediately post-intervention, and at 6-month follow-up. Analysis will use linear mixed-effects models to provide preliminary estimates of effect and estimate key parameters for a future definitive trial. The trial is embedded within a concurrent realist evaluation that includes qualitative methods to explore the mechanisms and contextual conditions shaping these outcomes.

**Discussion:** This study is the first pilot trial to evaluate a sexual violence primary prevention intervention in Australian secondary schools. In addition to the trial focused on sexual violence prevention outcomes, its integration with realist evaluation methodology will advance knowledge about how, when, and for whom these outcomes occur. The study findings will inform adaptability and scalability for secondary schools nationally and internationally.

**Trial registration:** The trial is prospectively registered with the Australian and New Zealand Clinical Trials Registry (reference: ACTRN12625000409471p). Registered 6 May 2025.

## Background

### Sexual violence and primary prevention

Sexual violence is a global public health concern and a gender-based issue arising from gender inequality and power imbalances [1, 2]. Over 1 in 5 (22%) women and 1 in 16 (6.1%) men in Australia have experienced sexual violence since the age of 15 [3]. In addition, over one in three young people in Australia indicate they have experienced unwanted sex during their life [4]. Research on trans and gender diverse people is still emerging, but evidence suggests they experience even higher rates of sexual violence than cisgender women [5]. Given its prevalence and harms, it is important to address the root causes of sexual violence, through primary prevention methods. Primary prevention ‘aim[s] to address the gendered drivers of violence against women, including the structures, norms and practices that maintain a gender unequal society’ [6]. School-based sex education, including relationship and sexuality education (RSE), and consent and respectful relationships education (CRRE) represent a promising area for primary prevention efforts. Their promise lies in their capacity to reach participants at a young age, before perpetration may have occurred and at a time in their lives where norms and values are being shaped [7, 8]. Schools are sites where young adolescents are socialised, and these early socialisation experiences shape attitudes and behavioural norms and can form developmental pathways that enable the perpetration of sexual violence [9]. Additionally, most young people spend a substantial amount of time at school, which presents a significant opportunity for intervention. Young people overwhelmingly believe RSE is an important part of the school curriculum [4]. However, they report being disappointed by the frequent emphasis in RSE on risk-reduction, and the conservative, heteronormative values underpinning much of this education [4]. Rather, young people are seeking RSE that is more forthright and imparts on them ‘practical skills to navigate sexual experiences’ including sexual practices and pleasure, and the inclusion of more diverse perspectives [10, 4].

Evidence suggests protective factors and enabling conditions can be built to prevent sexual violence, such as strengthening supportive relationships with parents and peers, and reducing social inequities, thereby enhancing social cohesion and collective wellbeing [11, 12]. Individual-level factors such as empathy, empowerment and interpersonal skills are also important protective mechanisms [13]. Good quality CRRE that nurtures the capacity of young people to build these protective factors in their lives is a key step in the primary prevention of sexual violence [14]. Indeed, school-based CRRE as a key strategy to reduce sexual violence has been recognised in the Australian Government’s *National Plan to End Violence Against Women and Children 2022-2032* [15].

Despite the significant harms of sexual violence, there is limited evidence of effective primary prevention interventions, that is, interventions designed to address the underlying cultural, structural, and normative conditions that give rise to sexual violence [16, 17]. While secondary schools can play an important role in primary prevention, a comprehensive review of sexual violence primary prevention interventions found no effective interventions in secondary school settings [16]. The review did identify eight promising interventions, including *Green Dot* [18], *Shifting Boundaries* [19], and *Men as Allies* [20], though their effectiveness varied and findings for behaviour change were inconsistent. Promising interventions shared some common approaches, such as a bystander component, gender sensitivity, and well-trained facilitators. They also tended to adopt ‘positive sexuality education’ and material ‘relevant to target audiences, such as age groups or cultural groups [17]. Importantly, secondary school-based interventions have generally not specifically targeted sexual violence and instead have been aimed more broadly at intimate partner or dating violence. Moreover, a vast majority of these interventions originate in the United States of America (USA), and therefore applicability to Australian schools is unknown. This gap is noticeable in the Australian context, where there is a dearth of peer-reviewed studies that have been conducted on primary prevention interventions to address sexual violence in secondary school settings [16].

Given the prevalence of sexual violence among young people, there is a critical need for effective primary prevention interventions targeting school settings in Australia. With specialised CRRE in secondary schools, they can become a setting where positive attitudes supportive of sexual violence prevention are shaped. To address this need, the Partners in Prevention of Sexual Violence project (PIPS) received funding from the Australian Department of Social Services to adapt and evaluate an existing program titled the ‘Schools Education Program’ (SEP) delivered by a community based sexual assault support service, to secondary schools. SEP takes a critical approach to CRRE that recognises the normative gendered and sexual cultures among young people attending school. The program responds to these cultures by critically engaging students in gender transformative discussions about consent and relationships that question deeply held notions of masculine entitlement and feminine passivity. One of the ways SEP does this is by challenging sexist myths about sexual violence known as ‘rape myths’, which refer to the notion that ‘rape can be prevented by verbal or physical resistance, and that women ‘ask for it’ via their actions’ [21]. In the three 90-minute student modules, SEP avoids individualising and pathologising language, instead framing primary prevention of sexual violence as best achieved through transforming social norms and structures.

This pilot cluster randomised controlled trial (cRCT) aims to test the primary hypothesis that students’ sexual violence knowledge, immediately after the SEP, will be higher for the treatment group than for the control group. Secondary hypotheses are that knowledge (after 6 months), attitudes (immediately and after 6 months) and behavioural intentions (immediately and after 6 months) will be higher for the treatment group than for the control group. Assessing changes in behavioural intentions rather than observed behaviour change was chosen, as behaviour change typically requires longitudinal follow-up over extended periods and substantially larger sample sizes to detect rare events that may not be feasible within the scope of a pilot trial. Behavioural intentions are established proximal predictors of future behaviour and are widely used as acceptable proxy measures in sexual violence prevention research, particularly in early-phase studies where the primary purpose is to establish feasibility and signal of effect [7, 22]. Demonstrating improvements in intentions to enact consent-related behaviours or intervene as prosocial bystanders provides meaningful preliminary evidence of program potential, while reserving definitive behavioural outcome assessment for a future fully powered trial should this pilot prove successful. This pilot trial is embedded within a broader realist evaluation seeking to understand how, when and for whom SEP generates its proposed outcomes, by theorising and testing explanations about implementation process and quality, as well as explanations about changes to participants’ intentions.

## Methods

### Study design

A two-arm, parallel pilot cRCT will be conducted among Year 9 students aged 13-15 years old in 12 secondary schools across one Australian state from 2025 to 2026. This pilot cRCT is the central component of a wider study that employs realist evaluation methodology. The realist evaluation will be conducted in parallel to develop and refine explanations of how, for whom and in what contexts SEP generates observed outcomes.

Given that SEP has not been previously evaluated in a controlled trial in Australia, this study will first be conducted as a pilot to test trial procedures, assess recruitment and retention rates, estimate parameters for a future definitive trial (e.g., intra-cluster correlation coefficients), and test the primary and secondary hypotheses. Randomisation will occur at the school level (cluster), rather than the individual level, because of the high risk of contamination in schools where students are routinely interacting. Ten of the 12 schools will be randomly allocated using covariate-constrained randomisation techniques to either the intervention (n=6) or a waitlist control (n=4); the two remaining schools are pre-assigned to the waitlist control group for logistical reasons. Six schools will receive the sexual violence primary prevention intervention (SEP) and six will be assigned to a waitlist control group. The control schools will not receive the program during the trial but, for ethical reasons, will be offered the intervention in late 2026, after study completion. The schedule of enrolment, interventions, and assessments is shown in the SPIRIT diagram (Figure 1).

**Figure 1:**
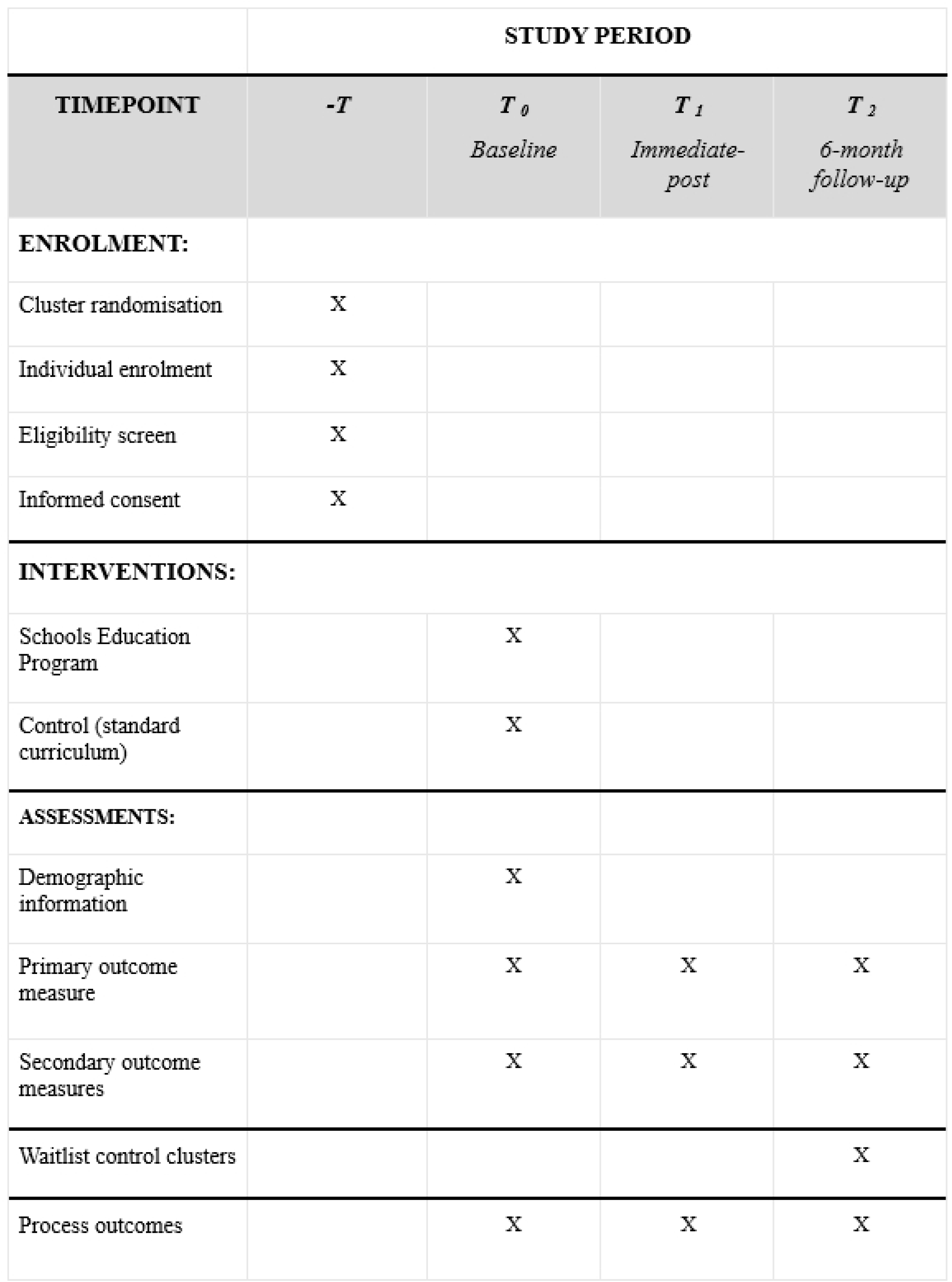
SPIRIT diagram of Schools Education Program trial

### Intervention

The SEP educational intervention aims to educate students on healthy sexual relationships and prevent sexual violence by enhancing student knowledge, attitudes and behavioural intentions. It is an age-appropriate intervention guided by the Australian government’s *Commonwealth Consent Policy Framework: Promoting healthy sexual relationships and consent among young people* [24]. SEP is also guided by the gendered drivers of violence outlined in the Our Watch document, *Change the Story* [1] and the societal-level and practical barriers to primary prevention of sexual violence set out in the National sexual violence prevention report and *Theory of Change* [21].

Year 9 students in the intervention arm will receive three 90-minute sessions of SEP facilitated by a specialised sexual violence service, in partnership with a state-based women’s legal service. The first session explores healthy relationships (communication, consent), sexual violence (myths, impacts), and gender equality (stereotypes, societal influences). In session two, a lawyer discusses legal consent, image-based abuse, stealthing, non-fatal strangulation, and resources for students seeking counselling, information or other supports for sexual violence related issues. In the final session, students learn consent skills, digital safety, and bystander strategies. This session explores topics such as how pornography shapes expectations of sexual intimacy, legal consequences of child abuse material, early warning signs of harmful behaviours, and safe tactics for intervening as a bystander. The intervention content is inclusive of a diverse range of sexual experiences, gender identities, and sexual orientations.

In addition to the student educational sessions, the intervention incorporates the recruitment and engagement of ‘School Student Champions’ within each participating school. The School Student Champions will meet regularly with SEP facilitators to provide student perspectives on the reception and impact of the education sessions. This component is designed to facilitate the diffusion of program messages throughout the broader student population, an evidence-informed strategy for normative change employed in analogous effective interventions [24, 25]. Additionally, members of the school staff and parents/caregivers will be invited to attend a 2-hour educational session on consent and respectful relationships to reinforce messaging. The program is informed by initial program theories about how (and when and for whom) SEP resources (e.g., gender-transformative content, legal knowledge, and bystander skill-building) are expected to shift students’ reasoning about relationships, consent, and social norms, thereby influencing knowledge, attitudes and behavioural intentions, particularly in school contexts where these messages are supported and reinforced. Initial program theories about program implementation (quality of program delivery, participant engagement, and school reinforcement of messages), are also included, though these are outside the scope of this protocol.

### Control

Schools in the control group will provide Year 9 students with the standard Australian state-based curriculum. This includes the Health and Physical Education (HPE) learning area, which contains a ‘Personal Care’ component, encouraging students to ‘reflect on and clarify their own values and attitudes’, to make ‘informed decisions about their personal health and wellbeing behaviours’ [Redacted]. The work requirement for this course is the development of a *Personal Wellbeing Plan* by each student, focused primarily on subjects unrelated to sexual violence, such as diet, nutrition, and fitness. So as not to disadvantage control schools, they will be waitlisted to receive SEP after the trial has concluded.

### Cluster and recruitment

The secondary schools will be recruited by the sexual assault service delivering the intervention, leveraging their existing networks to directly recruit eligible schools. Participant recruitment commenced on 18 August 2025 and is expected to conclude in Q2 2026. Data collection is expected to be completed in Q4 2026, with results expected in Q4 2027.

At enrolment, written informed consent will be obtained from both the parents/carers of student participants and from the student participants themselves. Consent will be provided via REDCap (Research Electronic Data Capture) software [26].

To be eligible, participants must meet the following criteria: students attending public secondary schools in the identified state of Australia; students in Year 9, aged approx. 13-15 years in these schools who consent to participate in the study and students whose parents or caregivers’ consent to their participation. The exclusion criteria include students attending non-government schools; students who do not consent to participate; students whose parents/caregivers do not consent to their participation, and students outside Year 9.

Twelve schools (clusters) have agreed to participate in this research. Due to logistical constraints in program delivery, two schools were unable to receive the intervention during the trial period and were therefore pre-assigned to the waitlist control group prior to randomisation of the remaining ten schools. For the randomised schools, a covariate-constrained randomisation procedure was used to allocate six to the intervention and four to the waitlist control group, ensuring covariate balance. The primary analysis will be based on the 10 schools that were randomly allocated. Data from the two pre-assigned schools will be included in the reporting of feasibility outcomes and may be used in supplementary sensitivity analyses, but they will be excluded from the primary hypothesis-testing analyses. To allocate the 10 randomised schools, 300,000 random allocations (6 intervention, 4 control) were simulated. For each allocation, the Mahalanobis distance was calculated based on the five following school covariates:

1. Year 9 student population (size)
2. Geographic region within the Australian state
3. Indigenous Australian student enrolment (percentage)
4. Index of Community, Socio-Educational Advantage (ICSEA)
5. Index of Relative Socio-Economic Disadvantage (IRSD)

The allocation with the smallest Mahalanobis distance (i.e., best overall balance across covariates between groups) was selected as the final assignment. These assignments will be communicated to the schools, and the baseline survey (T0) will be completed. Prior to T0, all trial team members, schools, and students will be blinded; after this point, all will be unblinded except the outcome’s assessor and members of the Data Monitoring Committee.

### Sample size estimation

Based on a pragmatic target of 12 participating schools (clusters) and an estimated average of 12 students per cluster (following adjustment for an anticipated 20% dropout rate), we aim to recruit approximately 192 students in total. In cases where the number of people for each school is not a whole number, it should be rounded up to the nearest greater integer when calculating the sample size for each cluster and accounting for the dropout rate. The number of participants recruited from each school will vary according to school student population size, ensuring proportional representation while maintaining the overall target sample size. This sample size is consistent with recommendations for pilot and feasibility studies, which typically do not require formal power calculations but rather focus on providing sufficient data to assess recruitment capability, retention, and the acceptability of trial procedures [27].

To provide context for future trial planning, a power calculation was performed using PASS 2024 (version 24.0.1). The sample size calculation above is based on the one-sided t-test, with degrees of freedom determined by the total number of subjects and a Type I error (α) of 0.025. Assumptions include a common subject-to-subject standard deviation of 1, an intra-cluster correlation coefficient of 0.026 [28], and a coefficient of variation for cluster sizes of 0.70926. At different attrition levels, if the average cluster size is 10 students per cluster instead of 12, then the power will be 0.76. If the average cluster size is 8, then the power will be 0.69; and if the average cluster size is 6, the power will be 0.58.

Participant retention will be promoted using reminders and vouchers. Students will be sent up to three automated reminders per survey. Students who participate in follow-up interviews (as part of the wider realist study) will be offered a $30 gift card. The structured school environment in which the program takes place will also serve as a mechanism to keep participants engaged throughout the study.

### Primary outcome

The primary outcome is the change in students’ sexual violence knowledge, measured immediately after the intervention. The author generated survey instrument comprises six items assessing participants’ knowledge. Respondents indicate their level of agreement with statements such as subjective norms (e.g., “I know what a healthy relationship looks like”), intrinsic empathy (e.g., “I try not to hurt other people’s feelings”), (e.g., “I believe someone when they tell me something bad has happened to them”), and (e.g., “Seeing other people upset makes me feel worries or sad”). Further, an item on pornography (e.g., “Porn shows what sex is really like”) and perceived prevalence of rape (e.g., “Unwanted sex is rare”) are included. Responses will be provided on a 7-point Likert scale (1 = Strongly disagree to 7 = Strongly agree).

### Secondary outcomes

The secondary outcomes are improvements in students’ attitudes toward gender equality and rape myths, and improvements in students’ behavioural intentions associated with consent and bystander intentions. Responses will be provided on a 7-point Likert scale (1 = Strongly disagree to 7 = Strongly agree). Student attitudes will be assessed using six items, including gendered beliefs (e.g., “Boys are better leaders than girls”), (e.g., “Girls are as smart as boys”), violence acceptance (e.g., “Sometimes violence is the only way to express your feelings”) and rape myth acceptance (e.g., “It is never a person’s fault if they are forced to have sex”), (e.g., “People think unwanted sex is a bigger deal than it really is”) and (e.g., “Girls often say ‘no’ to sex when they really mean ‘yes’”).

Lastly, there are four items assessing behavioural intentions. These include consent intentions (e.g., “I would ask for verbal consent to hug, kiss or touch someone”), (e.g., “I would listen if someone asked me to stop hugging, kissing or touching them, even if I still really wanted to do it”) and bystander intentions (e.g., I would tell a friend to stop if I heard them making a hurtful sexual joke about someone”) and (e.g., I would interrupt a friend who was hugging, kissing or touching someone who did not like it”). Further details on outcome measures are provided in the subsequent section.

### Data collection and assessments

Subsequently, the parents and carers of students receiving SEP, and students themselves, will give their informed consent to participate in the survey and/or interview. Participants will receive three quantitative assessments at baseline (T0), immediately-post intervention (T1), and again at 6-month follow-up (T2). The baseline survey will be completed by students before the commencement of the program in schools. Students will complete the surveys online via REDCap. The baseline survey will collect data on participant demographic and potential confounding variables, as well as hypothesised outcome variables. Immediate-post surveys will contain the same hypothesised outcome variables but exclude the demographic data. Six-month follow-up surveys will contain an additional item intended to collect self-reported data on which program sessions the students found most impactful.

Control and intervention group data will be collected concurrently to provide time-specific comparisons. The intervention group will receive staggered implementation due to facilitator scheduling constraints, and therefore, the control group data will be collected at the same time as the first wave of intervention group data to ensure alignment.

### Student surveys

Primary and secondary outcomes will be measured using a survey developed by the authors for this study. This survey comprises three subscales: knowledge (primary outcome, 6 items), attitudes (secondary outcome, 6 items), and behavioural intentions (secondary outcome, 4 items). The remaining six items collect demographic data from participants. Since previously used instruments are from USA-based studies, which did not capture the objectives of this study, and the language was not suitable for an Australian target population, the research team developed the survey for this study. A key objective of this pilot trial is to assess the internal consistency, acceptability, and preliminary psychometric properties of this newly developed instrument to inform its refinement for use in any subsequent definitive trial. While the sample size of 192 may limit the robustness of factor analytic techniques, preliminary psychometric assessment (e.g., Cronbach’s alpha for each subscale) will provide indicative data to guide refinement of the instrument. The survey is theoretically underpinned by existing validated measures from studies on gender-based violence, general violence, or sexual violence. These are the self-restraint – Weinberger Adjustment Inventory and the Basic Empathy Scale (BES) [29]; Attitude Toward Women Scale for Adolescents (AWSA) [30]; Couple Violence Scale (ACVS) [31]; Dating Self-Protection Against Rape Scale (DSPARS) [32]; VicHealth 2006 Community Attitudes to Violence Against Women Survey [33]; Bystander Attitude Scale-Revised (BAS-R) [34].

### Implementation outcomes

In line with Medical Research Council Guidelines [35] for complex interventions, a process evaluation has been incorporated in the realist evaluation. The integrated approach allows for the theorisation, testing, and refinement of implementation outcomes, such as reach, retention, and fidelity, alongside other program theories.

To assess these implementation outcomes, facilitators delivering the intervention will complete fidelity sheets after each student session, recording student attendance, deviations from planned content, and other challenges or patterns of implementation. Tracking of program participant engagement will also be captured through these sheets. Acceptability and sustainability will be explored via interviews with students, facilitators and other key stakeholders. The broader realist evaluation, while outside the scope of this paper, will use interviews and focus groups with students, school staff, facilitators, and stakeholders to investigate in-depth the mechanisms (participants’ responses to program resources) and contextual conditions that shape these trial outcomes. A central focus will be the mechanisms triggered within, and contexts shaping, the implementation process itself. Data on implementation process and quality from the process evaluation will serve as key evidence for developing and refining program theories about how, when, and for whom the implementation of SEP shapes its outcomes.

### Participant timeline

**Figure 2:**
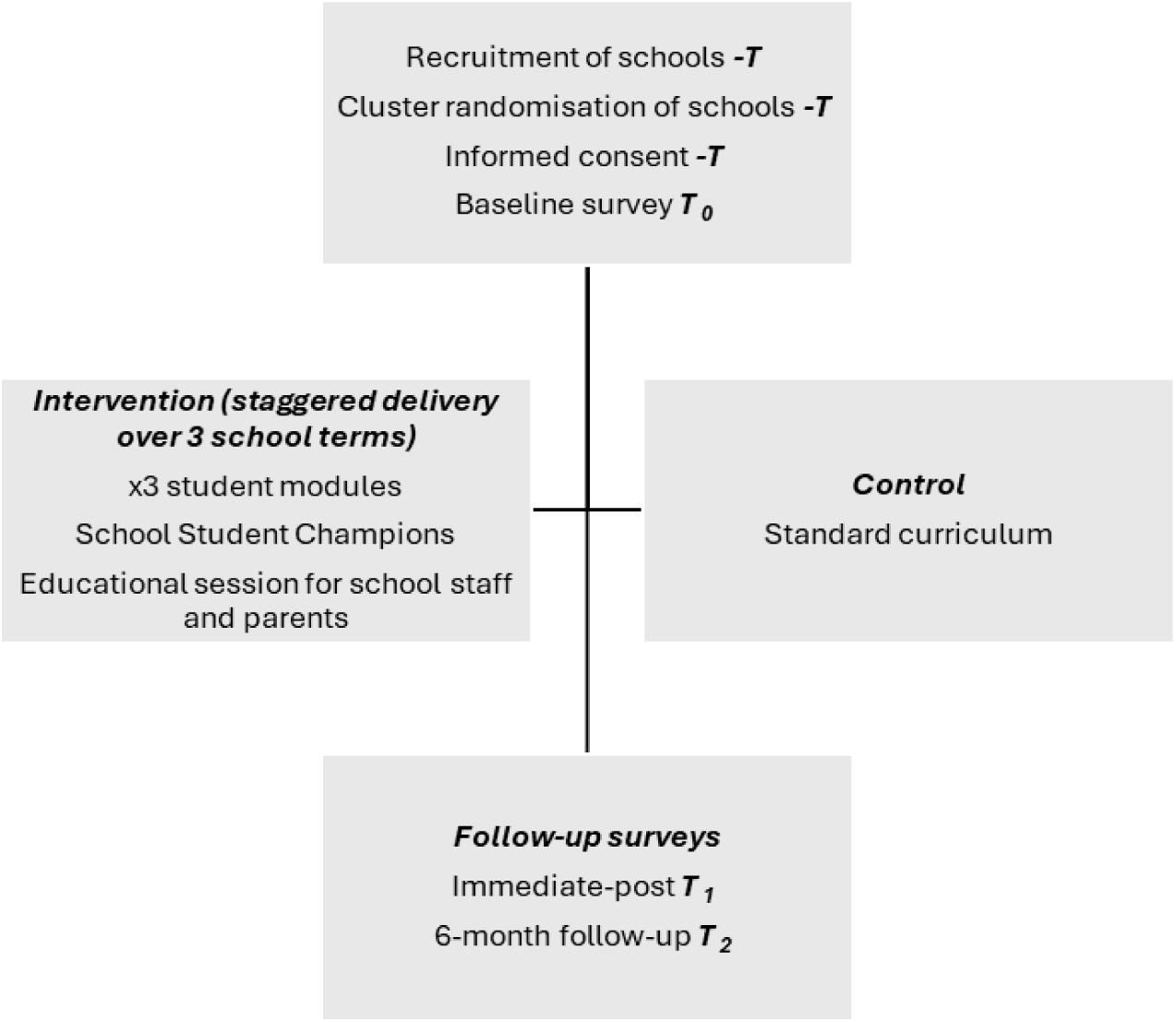
Flow diagram of participants throughout trial.

### Statistical analysis

Study results will be reported in accordance with the CONSORT extension for randomised pilot and feasibility trials, as well as the CONSORT guidelines for cluster randomised trials. The analysis will estimate intervention effects for the primary and secondary outcomes and report feasibility outcomes. The proportion of students completing baseline, immediate-post and six-month follow-up surveys will be reported. A descriptive statistical analysis will be used to summarise and describe the baseline student characteristics (categorical or continuous variables), such as age, gender, sexual orientation, Indigeneity, country of birth, language spoken at home, and self-reported learning disabilities. Descriptive analyses, including means and standard deviations for the primary and secondary outcomes, will be reported at baseline, immediately post-intervention, and at follow-up.

The primary outcome is the change in knowledge score, defined as the difference between post-intervention and baseline knowledge scores to test the primary confirmatory hypothesis that students’ sexual violence knowledge, immediately after the SEP, will be higher for the treatment group than for the control group. The main analyses will provide estimates of effect for this hypothesis and the secondary hypotheses. For the primary outcome, the analysis will estimate the mean change in student knowledge scores between the intervention and control groups using a linear mixed-effects model. The primary analysis will be conducted using a linear mixed-effects model, including a random intercept for cluster to account for intra-cluster correlation, a fixed effect for group allocation (intervention vs. control), and a fixed effect for baseline knowledge score to adjust for initial differences and to improve statistical efficiency. The model can be expressed as:

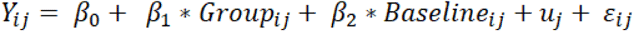

Where *i* indexes students and *j* indexes schools; *u*_*j*_ represents the random intercept for school to account for clustering of students within schools.

Additional covariates collected at both the student and cluster levels (e.g. student age, gender, country of birth, language spoken, and school-level covariates used in the randomisation procedure) will be included in the model further to explore their potential influence on the outcome. All effect estimates will be presented with confidence intervals and interpreted cautiously as preliminary estimates. These estimates will be used to inform sample size calculations and design parameters for a future definitive trial. The group effect will be tested using a one-sided hypothesis test with a significance level of α = 0.025. This threshold was selected to align with the a priori sample size calculation and to provide conservative control of the Type I error rate. All other tests will use a two-sided significance level of α = 0.05.

Secondary outcomes, including student sexual violence attitudes and behavioural intentions, will be analysed using the same linear mixed effects model approach as the primary outcome, accounting for clustering and adjusting for baseline measures where available. Furthermore, for both primary and secondary outcomes, all three time points (baseline, immediate post-intervention, and six-month follow-up) will be analysed using linear mixed-effects models that include fixed effects for group, time, and their interaction, as well as random intercepts for clusters and individuals. Baseline measures will also be included. An intention-to-treat analysis will be applied. Missing data will be assessed using Little’s MCAR test [36]. If non-significant, complete-case analysis will be used; if significant, multiple imputation will test robustness. Feasibility outcomes (e.g., recruitment and retention rates) will be reported descriptively.

### Realist evaluation

This pilot cRCT is embedded within a wider realist evaluation. The realist evaluation will integrate quantitative trial data with qualitative data to develop and refine explanations of how SEP works, for whom, and in which contexts, through analysis of context–mechanism–outcome relationships [37]. The full realist evaluation will be reported elsewhere.

### Ethical considerations

Human Research Ethics Committee approval was granted by La Trobe University (HEC25012) and the state’s Department for Education, Children and Young People. Adverse events will be monitored by the Data Monitoring Committee (DMC) and Trial Management Committee (TMC), and decisions made to terminate the trial if necessary. Schools will report adverse events, with serious events escalated immediately. Participation is voluntary, with parental opt-out available. A data management plan is to be conducted in accordance with ethics approval. Survey data will be non-identifiable; interview, focus group, and fidelity sheet data will be identifiable at collection. Sensitive information collected includes sexual orientation, country of birth, disability status, and Aboriginal and/or Torres Strait Islander identity. Indigenous Data Sovereignty principles apply.

Data are stored securely on La Trobe University’s Research DataSpace and REDCap, with access restricted to the research team. Audio recordings are deleted following transcription. The PIPS Chief Investigator is the data custodian. Data will be retained until participants turn 18 years, then for a further seven years. De-identified survey and fidelity sheet data will be retained indefinitely by the Department of Social Services. At study completion, data held by the research team will be securely retained by the Chief Investigator and permanently deleted from institutional platforms after the retention period.

## Discussion

This paper outlines the protocol for what is, to our knowledge, the first pilot trial of an intervention aimed at the primary prevention of sexual violence in Australian secondary schools. Grounded in evidence from promising school-based interventions abroad and underpinned by a national sexual violence prevention Theory of Change [21] and the Commonwealth Consent Policy Framework [23], this pilot study will test the primary hypothesis that students’ sexual violence knowledge, immediately after the SEP, will be higher for the treatment group than for the control group, and secondary hypotheses for knowledge after 6 months, attitudes (immediately and after 6 months) and behavioural intentions (immediately and after 6 months), while gathering data to determine whether a larger, fully powered definitive trial is warranted and feasible. Key uncertainties to be addressed include the willingness of schools to be randomised, the ability to recruit and retain adolescent participants over a six-month follow-up period, the suitability of the primary and secondary outcomes, the acceptability of the newly developed survey instrument, and the practical challenges of implementing SEP across diverse school contexts.

This pilot trial is distinct from, but embedded within, a broader realist evaluation. The realist study, to be reported separately, will also provide explanatory insights about program mechanisms and contexts, including those related to implementation quality, that shape the outcomes measured in this trial. By theorising the implementation process itself, the evaluation will identify the mechanisms triggered by different implementation approaches and the contexts that influence their effectiveness. This integrated approach will enable early identification of factors critical to intervention success, informing both program refinement and the design of a future definitive trial.

This study used a covariate-constrained randomisation approach in which the final allocation was selected from multiple candidate randomisations based on covariate balance. Although analyses adjusted for the covariates used in the allocation, this approach does not fully account for the selection process and may result in underestimated standard errors. Accordingly, findings should be interpreted with caution. Future studies may consider alternative allocation or inference methods that explicitly account for constrained randomisation.

If the intervention demonstrates preliminary evidence of effect and proves feasible to evaluate, this study will make significant contributions to the field of sexual violence primary prevention. To facilitate knowledge translation, study findings will be disseminated through peer-reviewed journals, conference presentations, and targeted reports. In doing so, the findings aim to contribute to community, governmental, and academic understanding of evidence-based strategies for the primary prevention of sexual violence.

## Data Availability

No datasets were generated or analysed during the current study. All relevant data from this study will be made available upon study completion.

## Trial status

The pilot cluster randomised trial of SEP began in Q3 2025. The results will be available by Q4 2027.

## Supplementary information

**Additional file 1:** Completed SPIRIT checklist for this SEP pilot cluster randomised controlled trial protocol article.

## Abbreviations

SEP: Schools Education Program
PICF: participant information and consent form
PIPS: Partners in Prevention of Sexual Violence project

## Acknowledgements

We thank Laurel House for their significant work developing the intervention, in particular Laurel House educators and the Lived Experience Advisory Panel for Young People. We would also like to thank the Partners in Prevention of Sexual Violence Lived Experience Think Tank for their guidance and lived expertise; the Partners in Prevention of Sexual Violence Project Advisory Group, particularly Vera Newman, for their feedback on the study design and methodology; and Hanorah Arundell for project support.

## Authors’ contributions

LH, JI, KF, FY, JT, GC, AB, & NH conceived of the trial and led the trial design, overall analysis plan (with lead support from FH), and the funding application. FH, LH, JI, KF, JT, GC, AB, NH, XL, JK, IM & SV will draft the manuscript. All authors will contribute to refinement of the study protocol and will read and approve the final manuscript.

## Dissemination policy

Trial results will be communicated to participants, relevant stakeholders and the public through a final report and publications which will be published on the Partners in Prevention of Sexual Violence website. The findings will be further disseminated through academic and practice conference presentations, the abstracts of which will be posted on the Partners in Prevention of Sexual Violence website.

## Declarations

## Ethics approval and consent to participate

This study was approved by the La Trobe University Human Research Ethics Committee (reference: HEC25012) and the [redacted] Department for Education, Children and Young People (reference: 2025-06). All study participants will be asked to provide informed consent to participate in data collection.

The Trial Monitoring Committee will guide the research and provide advice and support throughout the study. All members of the research team have Victorian issued Working with Children Checks (WWCC), with FH and JT possessing a Tasmanian issued Working with Vulnerable People Check.

## Consent for publication

Not applicable.

## Data availability statement

The de-identified quantitative dataset, code, and data dictionary will be provided to researchers by the Chief Investigator within 30 days of a request. Qualitative data are not available for sharing due to the risk of deductive disclosure given the sensitive nature of the topic and the small number of participating schools.

